# More prevalent, less deadly? Bayesian inference of the COVID19 Infection Fatality Ratio from mortality data

**DOI:** 10.1101/2020.04.19.20071811

**Authors:** G. W. Delius, B. J. Powell, M. A. Bees, G. W. A. Constable, N. J. MacKay, J. W. Pitchford

**Affiliations:** Department of Mathematics, University of York, York, UK, YO10 5DD; Department of Biology, University of York, York, UK, YO10 5DD

## Abstract

We use an established semi-mechanistic Bayesian hierarchical model of the COVID-19 pandemic [1], driven by European mortality data, to estimate the prevalence of immunity. We allow the infection-fatality ratio (IFR) to vary, adapt the model’s priors to better reflect emerging information, and re-evaluate the model fitting in the light of current mortality data. The results indicate that the IFR of COVID-19 may be an order of magnitude smaller than the current consensus, with the corollary that the virus is more prevalent than currently believed. These results emerge from a simple model and ought to be treated with caution. They emphasise the value of rapid community-scale antibody testing when this becomes available.

## 1 Introduction

A central question of the COVID-19 pandemic is whether herd immunity has been or is being developed to any useful extent. A reliable antibody test would rapidly elucidate the underlying state of immunity across a community. For the present, we need to try to infer the level of immunity from other available data.

Here our aim is to discover what the series of daily deaths (the most reliable of existing datasets for the European epidemic) tells us about the infection-fatality-ratio (IFR), the proportion of infections which results in death. Survival is assumed to result in immunity, which makes this ratio the crucial parameter linking deaths to the dynamic development of herd immunity.

We seek to understand IFR rather than the perhaps more commonly used CFR (case-fatality-ratio); the novelty of COVID-19 and the difficulties in its detection make the identification of ‘cases’ imprecise at the current time. In contrast, the definition of ‘infection’ is unambiguous – an infected individual is someone who can infect other susceptible individuals.

## 2 The Model

There are multiple factors that influence the evolution of the epidemic and whose relative importance are unknown. In [1] the COVID-19 Response Team at the MRC Centre for Global Infectious Disease Analysis at Imperial College propose a semi-mechanistic Bayesian hierarchical model to attempt to infer the impact of the various non-pharmaceutical interventions (self-isolation, social distancing, ban of public events, school closure and complete lockdown) from the time series of daily deaths in 11 European countries, later extended to 14 countries [2]. The timing of these interventions in the different countries is known, and this allows their impacts to be separated in the data. The model is a stochastic implementation of the classic SIR (susceptible-infected-removed) model structure in which individuals transition between these states over time. The results from the model of [1] can be viewed online at https://mrc-ide.github.io/covid19estimates/#/. We are grateful to the Imperial College COVID-19 Response Team for the exemplary accessibility and clarity of their work, including making their code available on GitHub at https://github.com/ImperialCollegeLondon/covid19model. The model assumes that each intervention reduces the basic reproductive number *R*_0_ for the virus by a constant factor to *R*_*t*_, starting on the day *t* that the intervention is imposed. This factor is assumed to be the same in all countries; only the date of the intervention differs between countries. This assumption allows the pooling of the data from all countries to extract the maximum amount of information. We will refer to this model as the ‘Flaxman model’ below.

The Flaxman model is used to provide a likelihood function for a Bayesian analysis of properties of the population and the virus simultaneously. Informally, a Bayesian analysis involves using the likelihood function to modify a set of current estimates for these properties encoded as a prior distribution. The location, shape and structure of these distributions are chosen to represent uncertainty in the current estimates. In accordance with Bayes’ rule for rational belief adjustment, the posterior distribution derived from the prior and the likelihood function expresses updated beliefs about the values of the model parameters given the data.

Direct, analytic calculation of the posterior distribution for the Flaxman model is intractable. Instead its properties are approximated from samples from the posterior. These are computed using Hamiltonian MCMC sampling as implemented by the statistical software Stan [3].

## 3 A reassessment of the Infection-Fatality Ratio for the Flaxman model

### 3.1 Proposed modification to the prior

The Flaxman model, as described in [2], specifies country-specific IFR values in several steps. Previous work from the Imperial group, specifically [4], is used to provide initial estimates that were originally computed alongside homogeneity assumptions across age-groups. These are then adjusted by factors understood to encode the effect of country-specific inter-generational mixing patterns. The adjusted estimates, denoted IFR_*m*_ with *m* indexing a country, are used to parameterize priors for the true country-specific IFR values, denoted 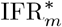:

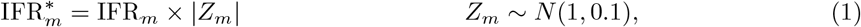

where the *Z*_*m*_ parameters allow for uncertainty around the IFR_*m*_ estimates and *N* (1, 0.1) denotes the normal distribution with mean one and standard deviation 0.1. We argue that the available mortality data can and does constrain the true 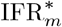 values away from the regions of greatest prior probability mass as specified by (1). It is possible to assimilate the extra information in the data by, for example, introducing an additional parameter *µ* that scales all of the country-specific IFR values. One convenient way to implement this is with the following prior specification:

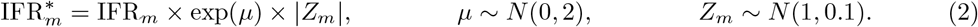

The normal prior on *µ* encodes a relaxation of the beliefs put forward in [2]. Specifically, it introduces the idea that the true IFR values may be systematically misspecified, and that the appropriate correction factor is *a priori* likely (with probability 0.95) to lie in the interval [exp(−1.96×2), exp(1.96×2)] ≈ [1/50, 50].

A concentration of posterior mass for *µ* away from zero provides quantitative support for the hypothesis that it is appropriate to revise the estimates arising from [2]. This is indeed the finding of our recent numerical experiments. More precisely, we find that for a range of priors, which accommodate a multiplicative adjustment to all countries simultaneously, posterior inferences strongly suggest parameter values consistent with revising down the initial IFR values by a factor of about ten.

The factor of *e*^*µ*^ applied to the IFR means that, in order to explain the same number of observed deaths, the number of infections will be higher by a factor *e*^−*µ*^. We therefore also scale the number of initial infections used in the Flaxman model by this factor. Simulations in which these numbers remained unscaled, although not presented here, lead us to near-identical posterior distributions.

### 3.2 Implications of the modified prior

The findings from our numerical experiments with the Flaxman model are usefully communicated in Figures 1 and 2.

**Figure 1:**
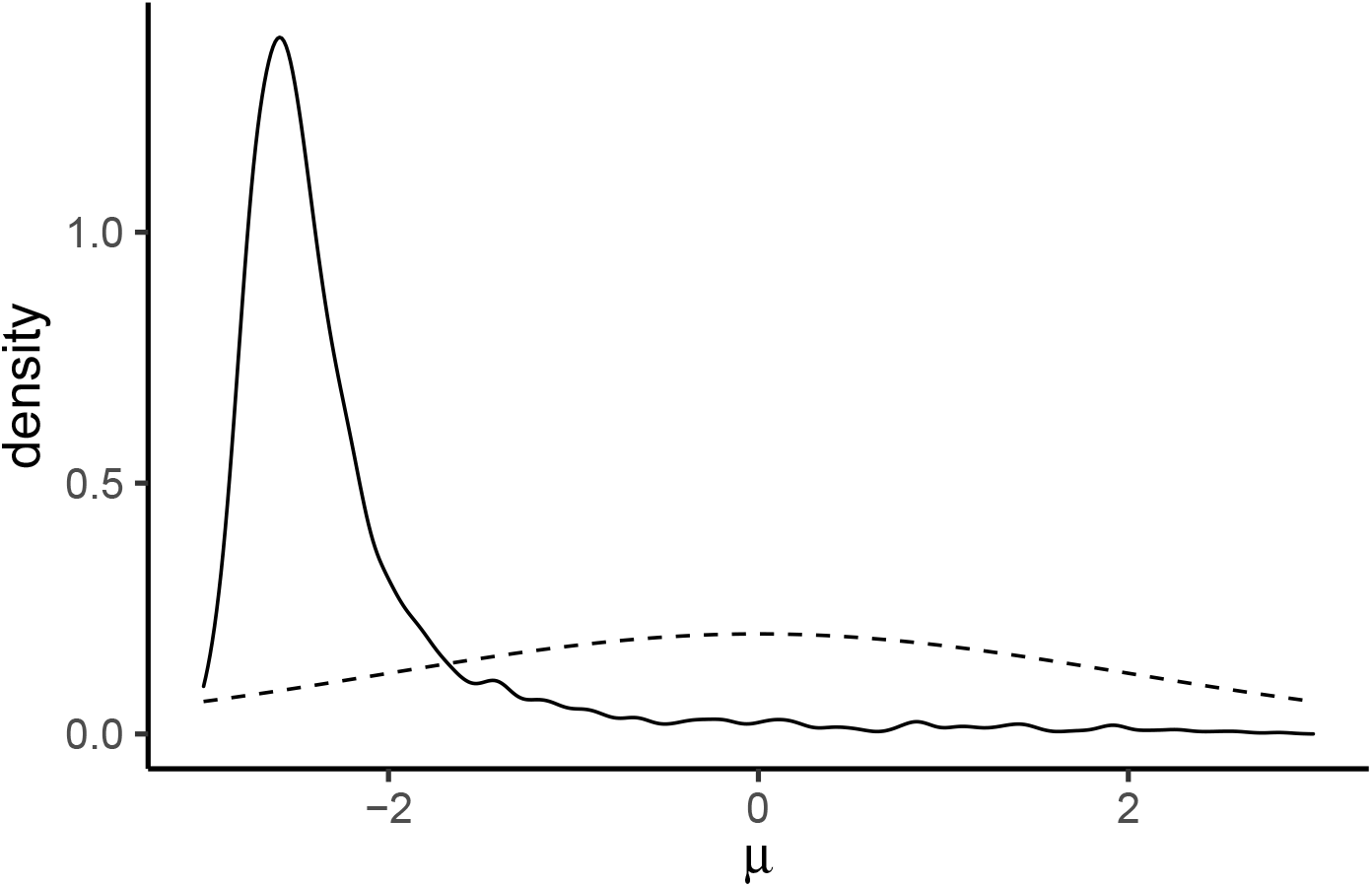
An approximate posterior (solid line) for the logged IFR adjustment parameter *µ*, whose role is defined in (2). The prior on *µ* is also illustrated here (dashed line). The concentration of posterior mass for *µ* away from zero provides quantitative support for the hypothesis that it is appropriate to revise the IFR values from [2], which used a prior that was tightly peaked around *µ* = 0.

**Figure 2:**
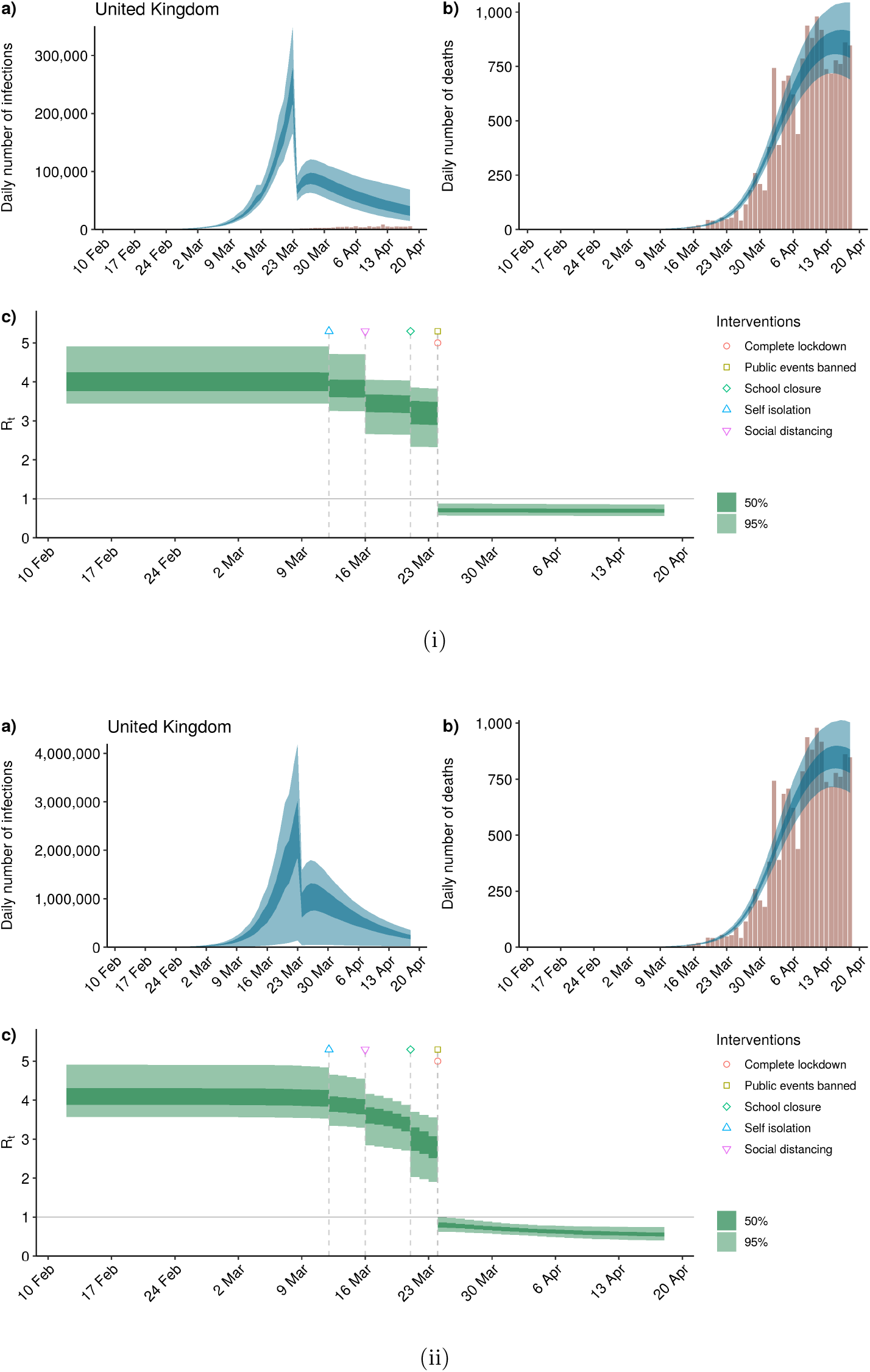
Daily infection, death and *R*_*t*_ estimates for the UK as produced by the code provided by [2]. Subfigures 2i and 2ii show the inferences consistent with priors (1) and (2), respectively. The nested blue regions illustrate 50% and 95% credible regions for daily infection and death rates. The heights of the vertical red bars correspond to observed deaths, which are the data that inform parameter estimates. The *R*_*t*_ estimates, whose likely values are also illustrated with 50% and 95% credible regions, are inferred via the equations of the Flaxman model.

In Figure 1 we plot an approximate posterior density function for the logged IFR parameter *µ*. We note in particular that 95% of its mass falls within the highest-posterior-density credible interval [−3.07,−0.53], corresponding to a downwards revision of the original 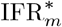 estimates by a factor between 0.0475 and 0.590. Additional results, computed using uniform priors on the adjustment parameter and which are not presented in the current document, lead to posteriors with very similar accumulations of mass. In this sense the modified model, with IFR values informed by the European death data, can be said to motivate significantly lower values than those inferred in [4].

In Subfigures 2i and 2ii we compare the daily infection rate, daily death rate and daily reproductive number (*R*_*t*_) estimates for the UK consistent with versions of the Flaxman model equipped with priors (1) and (2), respectively. The smaller 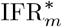 values, made accessible by the relaxed prior (2) but informed by the European data via the model, lead to correspondingly larger estimates for the number of daily infections over March and April. The death rates and the implied effects of interventions on *R*_0_ to produce *R*_*t*_, however, are affected to a much smaller degree. We note that the additional uncertainty introduced by prior (2) also leads to considerable additional uncertainty for these infection estimates.

Table 1 shows the (95%) credible intervals of possible fractions of the population in each country that have been infected, for our estimated IFR and for the fixed IFR of [1]. Its most salient feature is that, despite the apparently narrow posterior distribution of *µ* in Figure 1, the range is large: in the UK, anywhere between 2.3% and 71% may already have been infected – that is, between 1.6 million and 48 million people. The mean is around 30 million.

**Table 1:** Model estimates of the mean and 95% credible interval for the percentage of the population that is immune on 18 April 2020. Left column uses our posterior IFR; right column uses the fixed IFR of [1].

| Countries | Immunity with estimated IFR |  | Immunity with fixed IFR |  |
| --- | --- | --- | --- | --- |
| Austria | 10.09% | [0.38% – 19.55%] | 0.82% | [0.58% – 1.17%] |
| Belgium | 70.29% | [6.54% – 91.51%] | 12.90% | [8.28% – 19.40%] |
| Denmark | 11.62% | [0.47% – 22.62%] | 0.95% | [0.67% – 1.33%] |
| France | 43.44% | [1.99% – 69.71%] | 4.35% | [3.18% – 6.04%] |
| Germany | 12.05% | [0.43% – 24.72%] | 0.89% | [0.61% – 1.31%] |
| Greece | 1.29% | [0.05% – 2.37%] | 0.11% | [0.08% – 0.15%] |
| Italy | 43.98% | [2.09% – 71.10%] | 4.03% | [3.16% – 5.21%] |
| Netherlands | 38.25% | [1.71% – 66.12%] | 3.39% | [2.51% – 4.55%] |
| Norway | 6.12% | [0.21% – 12.42%] | 0.49% | [0.34% – 0.72%] |
| Portugal | 12.22% | [0.49% – 23.20%] | 1.00% | [0.72% – 1.39%] |
| Spain | 59.33% | [2.98% – 87.00%] | 6.05% | [4.57% – 7.99%] |
| Sweden | 61.38% | [4.76% – 90.31%] | 9.88% | [4.74% – 18.55%] |
| Switzerland | 22.81% | [0.93% – 41.22%] | 1.93% | [1.43% – 2.62%] |
| United Kingdom | 44.09% | [2.33% – 71.23%] | 4.37% | [3.22% – 5.90%] |

### 3.3 Modelling considerations

As explained above, the Flaxman model with a broader prior distribution for the IFR infers a posterior distribution for the IFR that suggests values that are up to an order of magnitude smaller than previously believed. We stress that this result relies on the assumption that the Flaxman model captures the essence of the epidemic adequately. We will now discuss various possible modifications of the model.

#### 3.3.1 Incorporating an inert class

We considered the possibility that a fraction *u* of the population is unable to be infected and unable to infect. (The mechanisms for such apparent ‘immunity’ are not considered here and may lie outside traditional definitions; nevertheless such a sub-population naturally contributes to herd immunity.) Simulations show that this has only a negligible effect on the IFR.

#### 3.3.2 Post-lockdown changes in contact structure

We are also concerned that the high apparent IFR may be at least in part an artefact of the very different networks of social structure that apply after lockdown and that cannot be accommodated by the Flaxman model, which assumes a continuous and well-mixed population. In reality, the postlockdown population contact network is likely to be highly modular, with the possibility of many infection-free subpopulations [5] (e.g. rural communities or small self-isolating households) that are effectively isolated from the main population over the timescale of available data.

The incorporation of an inert class, as in the previous subsection, could to some extent mimic the existence of a proportion of post-lockdown isolated subpopulations within the model. However, a crucial difference from the previous section is that the contribution to herd immunity provided by this population structure is reversible and would be lost on the release of social distancing measures.

#### 3.3.3 Implementation of lockdown

We considered the effect of introducing the impact of each intervention more gradually over one infectious period, so that the reproductive number *R*_*t*_ is no longer a step function. This removes the abrupt jumps in the estimates of the daily number of infections visible in Figure 2. Such a gradual decrease in *R*_*t*_ is more realistic, for example because after the lockdown individuals already infected may continue to infect other members of their household. Our simulations show that while this alters the relative effectiveness of the different interventions, it has only a negligible effect on the IFR.

## 4 Discussion and Conclusions

These results come from a simple model, authored by a leading group, in which the complexities of demography, and the diversities of an individual’s chance of, and response to, infection, are compressed into independent random variables which describe and link simple categories. This allows transparency, and clarity of interpretation, but sacrifices the detail offered by models with more complex transmission possibilities (e.g. [6]) and by data-intensive approaches (e.g. [7]). Naturally, such disparate approaches ought to be regarded as complementary and any conclusions treated with appropriate caution.

One very powerful aspect of this simple approach is that it considers only mortality. While it is true that there remain biases and imprecisions (figures may be under-reported; death *from* COVID-19 is not death *with* COVID-19), such inaccuracies are unlikely to explain the observed order-of-magnitude changes in IFR. We note that under-reporting by a constant ratio simply translates into the same ratio applied to the IFR, but does not affect the levels of herd immunity implied by Table1. Current European data may also confer an advantage over previous work based on data solely from China [4].

Our results suggest that, for European data at national scales, IFR estimates may be reduced by a large factor. A preliminary study [8] in Santa Clara County, California, estimates that COVID-19 seroprevalence exceeds the reported confirmed cases by a factor compatible with our analysis. Qualitatively similar results suggesting increased prevalence emerge also from Gangelt (Germany) [9]. In confined environments such as the Diamond Princess cruise ship [10] it is also possible that infection was more widespread than reported cases (but the unusual age distribution plays a crucial role in this case). This issue needs closer scrutiny as testing improves.

Our simple model suggests that the proportion of the population who cannot, or can no longer, be infected or infectious is larger than currently-accepted opinion. As we see from Table 1, the range of possibilities supported by the data is large, but the immunity level in the UK, for example, is probably much greater than current estimates, and a significant fraction of the population. Notice that this is not quite the same as saying that the prevalence of COVID-19 in the population is much higher than currently believed: recall that the model cannot discriminate well between the effects of IFR and of an inert class. The former will be directly tested with the advent of practicable community-scale antibody testing, while the latter, if it exists at all, is likely only to become clear gradually with a wide range of work. However, it is the sum of effects leading to herd immunity – whether initial, dynamically developing, or some combination of these – which is crucial.

We do not consider that our results have any immediate implications for policy in releasing lockdown or relaxing social distancing. Our short-term forecasts do not differ greatly from those of the Flaxman model – rather we emphasize that present data and near-future forecasts can be consistent with very different underlying IFR. Future policy decisions must necessarily embody a precautionary principle, carefully engineering an effective reproductive number which appropriately balances social and economic needs – a political decision. For example, to mitigate the reversal of effective herd immunity gains we might adopt a strategy of alternating relaxation of lockdown [11].

Our results suggest three possibilities. Either 1) the higher IFR estimated on the basis of early data and used in [1], while unlikely in the context of more recent fatality data, is correct, 2) the model requires further refinement, such as including age or network structure, or 3) the Flaxman model is sufficient, the data are reliable, and with high probability the IFR is much lower than has been the prevailing view, with herd immunity developing over a period of months.

Code for reproducing the results of this report and plots of the results for other European countries can be found at https://github.com/gustavdelius/covid19model/, where also further development is taking place.

## Data Availability

Code for reproducing the results of this report and plots of the results for other European countries can be found at https://github.com/gustavdelius/covid19model/

## 5 Acknowledgments

We are grateful to the University of York’s RAMP team for fruitful discussions, and to the COVID-19 Response Team of the MRC Centre for Global Infectious Disease Analysis at Imperial College for making their models and code publicly available.

